# Might first-hand experience of ill-health and economic hardship during the COVID-19 pandemic strengthen public support for vaccination and the reallocation of health sector funding towards health emergency preparedness in South Africa?

**DOI:** 10.1101/2021.12.05.21267315

**Authors:** R Mattes, K Dalal, H Rhoma, S Lambert, T De Wet, GTH Ellison

## Abstract

**Aims:** We examined whether first-hand experience of ill-health and economic hardship during the COVID-19 pandemic might strengthen public support for vaccination, and for the reallocation of health sector funding towards health emergency preparedness in South Africa – a country in which high rates of vaccine hesitancy go hand in hand with widespread discontent regarding public service delivery.

**Methods:** Using data from 1,600 South African respondents who were surveyed during 2021 for the Eighth Round of Afrobarometer (AB-R8), discrete measures of household- and individual-level sociodemographic and economic factors were generated to permit confounder-adjusted analyses of probabilistic causal relationships between self-reported measures of: personal/household COVID-19 illness and job/income/business loss as a result of COVID-19; and the likelihood that respondents would accept a (government-approved) COVID-19 vaccine, or support the reallocation of health sector funding towards health emergency preparedness.

**Findings:** There was little evidence that personal/household experience of COVID-19 illness was associated with the likelihood that respondents would (or would not) accept a (government-approved) COVID-19 vaccine (OR: 0.96; 95%CI: 0.72,1.28); or that these respondents would (or would not) support the reallocation of health sector funding towards health emergency preparedness (OR: 0.95; 95%CI: 0.71,1.26), even after adjustment for individual- or household-level sociodemographic and economic covariates considered likely confounders. There was similarly little evidence that personal/household experience of job/income/business loss as a result of the COVID-19 pandemic was associated with support for the reallocation of health sector resources for emergency preparedness (OR: 1.02; 95%CI: 0.80,1.30); again, even after adjustment for potential confounders.

However, respondents who reported that they or someone in their household had lost their job/income/or business as a result of the COVID-19 pandemic had only around half the odds of reporting that they would accept a (government-approved) COVID-19 vaccine (OR: 0.60; 95%CI: 0.47,0.77) – and this finding, like the others in these analyses, was largely unaffected by the inclusion/exclusion of covariates considered susceptible to change following the onset of the COVID-19 pandemic (i.e. those covariates potentially operating as colliders rather than genuine confounders).

**Conclusions:** These findings suggest that – despite the postulated ‘experiential dividend’ of COVID-19 illness (i.e. its expected impact on vaccine hesitancy and support for the reallocation of health sector resources for health emergency preparedness) – no such ‘dividend’ was observed in this broadly representative sample of South African adults. Indeed, job/income/business loss (and associated economic hardship) also had little effect on support for the reallocation of health sector resources for health emergency preparedness; yet this was somewhat paradoxically associated with a much lower odds of vaccine acceptance – paradoxically, since vaccination has been widely viewed as a pragmatic (if somewhat neoliberal) intervention to protect economic activity. However, these findings might simply reflect inadequate confounder adjustment for preceding and entrenched attitudes towards vaccination amongst those South Africans who are also most vulnerable to job/income/business loss as a result of the COVID-19 pandemic. Protecting the livelihood *and* health of such individuals and households is likely to remain a substantial challenge and key priority for future emergencies in which economic activity is compromised.

**Aim:** The aim of the present study was to examine whether first-hand experience of ill-health and economic hardship during the COVID-19 pandemic might have strengthened public support for vaccination, and for the reallocation of health sector funding towards health emergency preparedness in South Africa – a country in which high rates of vaccine hesitancy go hand in hand with widespread discontent regarding public service delivery. To this end we drew on data generated by the eighth round of household surveys undertaken by Afrobarometer (AB-R8) which interviewed 1,600 respondents across South Africa during 2021 – more than a year after the country’s first confirmed case of COVID-19 on 5 March 2020.

## Methods

### Analytical design

To examine the putative causal relationships between each of the two specified exposures (a respondent and/or household member becoming ill with COVID-19; and/or losing their job/income/business as a result of COVID-19) and each of the two specified outcomes (willingness to accept a [government-approved] vaccine; and/or support for the reallocation of health sector resources to health emergency preparedness) we generated a causal path diagram (in the form of a directed acyclic graph [DAG]; see Figure S1, below) in which both of the more phenomenological exposures were assumed to have preceded the two more opinion-based outcomes. Temporal logic was then applied to identify sociodemographic and economic items measured by the AB-R8 survey instrument that were considered likely to have preceded both exposures (and therefore also both outcomes). In the absence of substantive evidence to the contrary, all such variables can be assumed to act as potential (or at the very least, ‘candidate’) confounders – i.e. as *probabilistic* causes of the specified exposures and outcomes that would require statistical adjustment in analyses to estimate each of the focal relationships examined, in order to mitigate the risk of confounding bias.

While this approach to causal inference using observational (i.e. non-experimental) data helps to reduce the impact of bias from *measured* confounders (by ensuring analyses condition thereon through sampling, stratification or – as in this study – statistical adjustment), the estimates generated are still likely to be biased as a result of: measurement error (i.e. residual confounding); a failure to adjust for unmeasured (latent) confounders (i.e. unadjusted confounding); and the misclassification of (and inappropriate adjustment for) mediators or consequences of the outcome as potential confounders (thereby generating collider bias).

In an effort to reduce residual confounding, we carefully examined the responses provided to each of the items selected as ‘candidate’ confounders to eliminate any measurement-related error generated by respondents with missing data values, and by overlapping/indiscrete answer options/categories – the first through case-wise deletion of respondents with (any) missing data; and the second through re-categorisation; though both of which would have nonetheless introduced alternative sources of bias and imprecision (particularly through selection bias and a loss of information, respectively).

There was less scope to address unadjusted confounding in the design of our analyses, not least given the finite number of items in the AB-R8 survey instrument and its emphasis on self-reported, opinion-based items that are challenging to interpret as phenomenological events (or crystallising processes) amenable to temporal positioning with respect to any given focal relationship.

Nonetheless, to acknowledge the potential role that such characteristics might play as unacknowledged or unmeasured confounders, we included four of the innumerable possible sets of unmeasured confounders within our theoretical causal path diagram (see Figure S1, below) to emphasise the (unadjusted confounding) bias they might bring to any estimates of the focal relationships examined in the present study.

Finally, since some of the ‘candidate’ confounders selected for adjustment comprised features of respondents or households that were subject to change over time (i.e. ‘time-variant’ variables), we sought to address uncertainty regarding precisely when these characteristics might have crystallised (i.e. as/when measured by the AB-R8 survey) by conducting sensitivity analyses with covariate adjustment sets containing ‘candidate’ confounders considered more vs. less likely to have themselves been affected by COVID-19 – the former including: respondent employment status; and respondent/household assets as indicators of any fluidity in socioeconomic position.

Notwithstanding our efforts to address these three potential sources of bias in the estimation of focal relationships from observational/non-experimental data (namely; residual confounding; unadjusted confounding; and colliders mis-specified as confounders), it is important to stress that these efforts are very unlikely to have been completely successful. Similarly, because incompletely representative sampling – which is common to most surveys involving relatively small samples of consenting participants (such as the AB-R8 survey) – can invoke endogenous selection bias (a form of ‘collider bias’), even carefully theorised causal path diagrams and the careful selection, measurement and adjustment for confounders may not eliminate the risk of generating biased causal estimates of focal relationships from analyses of observational data. For these reasons, the findings generated by the present study remain speculative and warrant careful examination, replication and further exploration.

### Selection of exposure, outcome and ‘candidate’ confounder variables

The two exposures and two outcomes of interest examined in the present study were derived from four items included in the supplementary (COVID-19) module which was inserted into the AB-R8 Questionnaire for those countries (including South Africa) where data collection had been suspended or postponed as a result of the emerging pandemic. The two items used to generate each of the exposures of interest asked respondents whether “*you personally or any other member of your household have been affected in any of the following ways by the COVID-19 pandemic:*” and listed two domains of effect, namely: “*Became ill with COVID-19*” and “*Temporarily or permanently lost a job, business or primary source of income*” for each of which the response options available were: Yes, No or Don’t Know. Likewise, the two items used to generate each of the outcomes of interest used the following item wording and answer formats to explore: first, the likelihood that respondents’ would accept a vaccination against COVID-19 (were this to become available) – “*If a vaccine for COVID-19 becomes available and the government says it is safe, how likely are you to try to get vaccinated?*” [Very un/likely, Somewhat un/likely, Refused, Not Applicable Don’t know]; and second, whether respondents agreed/disagreed that more government funding was necessary to prepare for health emergencies – “*Do you agree or disagree with the following statement: Our government needs to invest more of our health resources in special preparations to respond to health emergencies like COVID-19, even if it means fewer resources are available for other health services?*” [Strongly dis/agree, Dis/agree, Neither agree nor disagree, Refused, Not Applicable, Don’t know]. The distribution of responses to each of these exposures and outcomes was carefully examined to generate binary categorical variables that optimised the distribution of responses with the conceptual integrity of the answers that each provided.

The covariates identified as plausible ‘candidate’ confounders from amongst those items included in the AB-R8 survey instrument focussed primarily on phenomenological characteristics that were least likely to be vulnerable to reporting bias or to have changed substantively as a result of the COVID-19 pandemic (i.e. to have ‘re-crystallised’ following the impact of illness or job/income/business loss). While this meant that a large proportion of the (opinion-based) items included in the AB-R8 survey instrument were discounted as suitable for use as measures of ‘candidate’ confounders, there were a sizeable number of more phenomenological items relevant to the sociodemographic characteristics of the respondent (i.e. age, gender, race/ethnicity) and household (i.e. primary language spoken in the home), and even to the socioeconomic position of both respondent (i.e. educational attainment) and household (i.e. type of dwelling and household utilities, services and amenities) that were considered unlikely to have changed in the 12-18 months following the onset of the COVID-19 pandemic. While responses to items on these characteristics were therefore considered ‘time-invariant’ (and to have occurred – or crystallised as/when reported in the AB-R8 questionnaire – *prior* to either of the present study’s specified exposures), there were a number of additional phenomenological criteria (including: respondent employment and respondent/household ownership of 6 specific assets) that were likely to have been more vulnerable to the impacts of the pandemic, and were therefore considered ‘time-variant’ (and therefore to have occurred – or crystallised as/when reported in the AB-R8 questionnaire – *after* the present study’s specified exposures). To address the risk that the latter might not constitute genuine confounders (but instead might act as mediator-colliders between the exposures and outcomes examined), we undertook sensitivity analyses involving two sets of multivariable logistic regression models in the first of which only time-invariant covariates were included as presumed confounders in the covariate adjustment sets used, and in the second of which both time-invariant and time-variant covariates were included (see *Statistical analyses* and *Results: Multivariable statistical analyses*, below). Prior to these analyses, the distribution of responses to all of the items selected as ‘candidate’ confounders was examined prior to re-categorisation to facilitate analysis and interpretation.

### Statistical analyses

Standard descriptive statistics (frequencies with percentages for categorical variables, and medians with inter-quartile ranges for continuous variables) were used to summarise the responses obtained for each of the two exposures, two outcomes and 22 covariates examined in the present study.

Respondents who were ineligible/unable/unwilling to answer (or did not know the answer to) any of the questionnaire items required to generate these data were excluded from the sub-sample of respondents subsequently included in the complete case analyses that followed. These analyses involved multivariable logistic regression models designed with reference to the theoretical causal path diagram described earlier (see Figure S1) in which the postulated temporal sequence of, and probabilistic causal relationships between each of these variables had been used to select covariates likely to have acted as potential confounders for the four focal relationships examined (i.e. between respondent/household COVID-19 illness and/or job/income/business loss; and vaccine acceptance and/or support for the reallocation of health sector resources to health emergency preparedness). Sensitivity analyses were undertaken to compare the impact of adjusting for time-invariate, and time-variant ‘candidate’ confounders – the former considered likely to be genuine confounders, the latter considered at risk of being mis-specified mediator-colliders.

## Results

### Sample characteristics

In preparation for analysis, the distribution of each of the specified exposures, outcomes and ‘candidate’ confounder covariates was carefully examined to facilitate their re-categorisation into coherent categorical and ordinal variables. This included reducing each of the exposures and outcomes to binary variables to permit analysis using logistic regression analysis, in which categories selected were determined at, or as close as possible to, the median value where appropriate.

These re-categorised variables have been summarised in Table S1 (below) which indicates that 1,309 respondents provided complete data on all 26 variables, while almost a fifth (291; 18.2%) had provided responses to one or more of the survey items (e.g. Don’t know, Refused, or Not applicable) that resulted in missing data values. Given the risk of endogenous selection bias (a form of collider bias;) in analyses that seek causal inference from unrepresentative samples – a risk that can already be high in any population surveys that are dependent on fallible sampling techniques (such as household surveys) – the distribution of responses obtained from participants providing complete data on all 26 variables was compared to those of participants who had not (see column 2 of Table S1). This comparison provided some reassurance that respondents who provided complete data on all 26 variables (the ‘complete case [sub]sample’) had sociodemographic and economic characteristics (both individual- and household-level) that were broadly comparable to those who had not. However, there were substantial differences in the distribution of race/ethnicity, such that only 4.2% of the complete case sample were classified by interviewers as ‘South or East Asian’, compared to more than twice as many of those who provided incomplete data (10.0%). At the same time, substantially fewer of the latter reported that they, or someone in their household, had been affected by COVID-19 illness (13.3% vs. 20.3%) or had lost their income/job/business as a result of COVID-19 (28.5% vs. 34.6%), while vaccine acceptance was substantively higher amongst respondents in the complete case sample (46.2%) when compared to those with any missing data (38.5%).

Notwithstanding these differences, and the risk of collider bias they might entail, the analyses that follow rely solely on those 1,309 respondents for whom data were available on all 26 of the variables examined and, as such, the results of these complete case analyses need to be interpreted with caution from a causal inference perspective. This (sub)sample of adult respondents comprised a similar number of men and women, with a median age of approximately 50 (range: 18-90). Most (70.2%) were classified as ‘Black/African’, with far fewer classified as ‘Coloured/mixed race’ (15.5%), ‘White/European’ (10.1%) or ‘South/East Asian’ (4.2%). Most (76.4%) spoke a non-European language (and predominantly an African language) at home; and although 61.5% had completed secondary education (or above), only around a third (37.2%) reported they had current employment that paid a cash income, and around a third of these (34.4%) were only employed part-time (see Table S1). As such, the complete case sample of South African respondents included in the analyses that follow are characterised by high levels of underemployment, and this is likely to have a substantial bearing on the proportion who reported that they, or someone in their household, had become ill with COVID-19 (20.3%) or had (temporarily/permanently) lost their income/job/business as a result of COVID-19 (34.6%) – particularly if, as seems likely, a substantial proportion of those who reported that they were unemployed at the time Round 8 of the Afrobarometer surveys took place had lost their employment as a result of COVID-19. Under such circumstances, this employment variable (and its associated impact on the assets owned by respondents and their household) may constitute a *consequence* of COVID-19 rather than a potential (preceding) *determinant* of the two specified exposures examined in the present study.

### Multivariable statistical analyses

To address the possibility that individual-level socioeconomic characteristics (i.e. employment and assets) might constitute a consequence (as opposed to a determinant) of the two specified exposures (COVID-19-related illness and income/job/business loss) – and might therefore constitute a mediator (and therefore a potential collider) rather than a genuine confounder in the causal relationships between COVID-19 illness, income/job/business loss and each of the two specified outcomes of interest (vaccine acceptance and support for the reallocation of health sector resources to health emergency preparedness) – three separate logistic regression models were used for each of the four focal relationships examined (see Table S2, below). The first of these models adjusted for none of the ‘candidate’ covariates considered possible confounders; the second adjusted only those individual- and household-level sociodemographic and economic covariates considered likely to have occurred/crystallised *before* the onset of the COVID-19 pandemic; and the third also adjusted for employment and (individual- and household-level) assets – i.e. time-variant characteristics that might have been directly affected by the COVID-19 pandemic and associated restrictions on economic activity.

These models indicated that all of the estimated relationships examined (i.e. between each of the two specified exposures and each of the two specified outcomes) were somewhat attenuated following the adjustment for covariates considered potential confounders, but that the relationships were not substantively altered following adjustment for those covariates believed to have crystallised *before* the onset of the COVID-19 pandemic (i.e. Model 2 & 5) or those covariates considered susceptible to change thereafter (i.e. those potentially operating as mediators rather than confounders in the focal relationships examined; Model 3 & 6). On the basis of these analyses, there was little evidence that personal/household experience of COVID-19 illness was associated with the likelihood that respondents would (or would not) accept a (government-approved) COVID-19 vaccine (adjusted OR: 0.96; 95%CI: 0.72,1.28; see Model 2, Table S2); or that these respondents would (or would not) support the reallocation of health sector funding towards health emergency preparedness (adjusted OR: 0.95; 95%CI: 0.71,1.26; see Model 5, Table S2). There was similarly little evidence that personal/household experience of job/income/business loss following the onset of the COVID-19 pandemic was associated with support for the reallocation of health sector resources for emergency preparedness (adjusted OR: 1.02; 95%CI: 0.80,1.30; see Model 5, Table S2). However, respondents who reported that they or someone in their household had lost their job/income/or business as a result of the COVID-19 pandemic had only around half the odds of reporting that they would accept a (government-approved) COVID-19 vaccine (OR: 0.60; 95%CI: 0.47,0.77; see Model 2, Table S2). This finding, like the others in these analyses, was largely unaffected (and only somewhat attenuated) following the inclusion/exclusion of covariates considered susceptible to change following the onset of the COVID-19 pandemic (i.e. those covariates potentially operating as colliders rather than genuine confounders; compare Models 2 and 3, and Models 5 and 6, Table S2).

## Discussion and conclusion

Despite the postulated ‘experiential dividend’ of COVID-19 illness (i.e. its expected impact on vaccine acceptance and support for the reallocation of health sector resources for health emergency preparedness), no such ‘dividend’ was observed in this broadly representative sample of South African adults. Indeed, job/income/business loss (and associated economic hardship) also had little effect on support for the reallocation of health sector resources for health emergency preparedness; yet this was (somewhat paradoxically) associated with a much lower odds of vaccine acceptance – paradoxically, since vaccination has been widely viewed as a pragmatic (if somewhat neoliberal) intervention to protect economic activity under such exigencies.

However, these findings might simply reflect inadequate confounder adjustment for preceding and entrenched attitudes towards vaccination amongst those South Africans who are also most vulnerable to job/income/business loss as a result of the COVID-19 pandemic. Levels of distrust in government, and levels of discontent with the slow pace of service delivery, run high in South Africa and precede the COVID-19 pandemic. While both are strongly associated with sociodemographic characteristics (particularly age, gender and race/ethnicity) that remain powerful and pernicious structural determinants of socioeconomic power and disadvantage more than 25 years after the end of apartheid, it is plausible that the powerful source of potential confounding associated with such deep-seated distrust of, and disaffection with, government and public services might not have been adequately addressed simply through adjustment for the sociodemographic and economic covariates included in Models 2 and 5, or Models 3 and 6 of the present study (see Table S2).

Further research is required to substantiate this hypothesis. In the interim, protecting the livelihood *and* health of South Africa’s most vulnerable (and disaffected) individuals and households is likely to remain a substantial challenge and key priority for future emergencies in which economic activity is compromised – particularly as long as distrust of the government and dissatisfaction with public services accompanies the high rates of under-employment, material deprivation and poor education evident from the AB-R8 survey.

## Data Availability

All data produced are available online at: https://afrobarometer.org/data

## Supplementary material

### Part 1: Exposure and outcome selection and (re)categorisation from items included in the supplementary COVID-19 module used in the postponed Round 8 of the Afrobarometer survey in South Africa, *following* the onset of the COVID-19 pandemic

#### 1.1 Exposures

*First-hand experience of illness* [COVID-19ill_dv]

| **AB_R8-COV2 Please tell me if you personally or any other member of your household have been affected in any of the following ways by the COVID-19 pandemic... Became ill with COVID-19** | Original - Yes |  | Re-categorised categorical - Yes |  |  |
| --- | --- | --- | --- | --- | --- |
|  | n | % | Category | n | % |
| Yes (1) | 284 | 17.8 | Yes (1) | 284 | 17.8 |
| No (2) | 1160 | 72.5 | No (0) | 1160 | 72.5 |
| Not applicable (7) | 150 | 9.4 | Not applicable, Refused or Don't know (.) | 156 | 9.8 |
| Refused (8) | 3 | 0.2 |  |  |  |
| Don't know (9) | 3 | 0.2 |  |  |  |

*First-hand experience of job/business/income loss* [COVID-19job_dv]

| **AB_R8-COV3 Please tell me if you personally or any other member of your household have been affected in any of the following ways by the COVID-19 pandemic... Temporarily or permanently lost a job, business or primary source of income** | Original - Yes |  | Re-categorised categorical - Yes |  |  |
| --- | --- | --- | --- | --- | --- |
|  | n | % | Category | n | % |
| Yes (1) | 492 | 30.8 | Yes (1) | 492 | 30.8 |
| No (2) | 954 | 59.6 | No (0) | 954 | 59.6 |
| Not applicable (7) | 150 | 9.4 | Not applicable, Refused or Don't know (.) | 154 | 9.7 |
| Refused (8) | 1 | 0.1 |  |  |  |
| Don't know (9) | 3 | 0.2 |  |  |  |

#### 1.2 Outcomes

*Vaccine hesitancy* [vaccine_dv]

| **AB_R8-COV17 If a vaccine for COVID-19 becomes available and the government says it is safe, how likely are you to try to get vaccinated?** | Original - Yes |  | Re-categorised categorical - Yes |  |  |
| --- | --- | --- | --- | --- | --- |
|  | n | % | Category | n | % |
| Very unlikely (1) | 598 | 37.4 | Unlikely (0) | 771 | 48.2 |
| Somewhat unlikely (2) | 173 | 10.8 |  |  |  |
| Somewhat likely (3) | 267 | 16.7 | Likely (1) | 647 | 40.5 |
| Very likely (4) | 380 | 23.8 |  |  |  |
| Not applicable (7) | 150 | 9.4 | Not applicable, Refused or Don't know (.) | 182 | 11.4 |
| Refused (8) | 2 | 0.1 |  |  |  |
| Don't know (9) | 30 | 1.9 |  |  |  |

*Health emergency resource allocation* [emergency_dv]

| **AB_R8-COV20 Do you agree or disagree with the following statement: Our government needs to invest more of our health resources in special preparations to respond to health emergencies like COVID-19, even if it means fewer resources are available for other health services?** | Original - Yes |  | Re-categorised categorical - Yes |  |  |
| --- | --- | --- | --- | --- | --- |
|  | n | % | Category | n | % |
| Strongly disagree (1) | 182 | 11.4 | Less than agree (0) | 576 | 36.0 |
| Disagree (2) | 224 | 14.0 |  |  |  |
| Neither agree nor disagree (3) | 170 | 10.6 |  |  |  |
| Agree (4) | 436 | 27.3 | Agree or strongly agree (1) | 821 | 51.4 |
| Strongly agree (5) | 385 | 24.1 |  |  |  |
| Not applicable (7) | 150 | 9.4 | Not applicable, Refused or Don't know (.) | 203 | 12.7 |
| Refused (8) | 3 | 0.2 |  |  |  |
| Don't know (9) | 50 | 3.1 |  |  |  |

### Part 2: Confounder selection/(re)categorisation from items included in the original instrument used in the postponed Round 8 of the Afrobarometer survey in South Africa (*following* the onset of the COVID-19 pandemic)

To capture multiple dimensions of the many potential confounders likely to have influenced the estimated relationships between each of the exposures and outcomes specified in the present study (see Figure 1 in the main body of the manuscript), the items included in the instrument used during Round 8 of the Afrobarometer surveys (ABR8Q1-ABR8Q123) were subjected to repeated close-reading to identify those covering key preceding sociodemographic and economic characteristics of survey respondents (i.e. characteristics capable of acting as potential confounders when examining the focal relationships specified in the present study). The items selected have been listed below, alongside the frequency of responses to each item’s answer categories, and any re-categorisation applied (whether as categorical or ordinal variables) to facilitate analysis and interpretation.

#### 2.1 Sociodemographic characteristics

*Respondent Age* [age_dv]

| AB_R8-Q1. How old are you? | Original |  |  | Re-categorised ordinal |  |  |
| --- | --- | --- | --- | --- | --- | --- |
|  | Mean | Min | Max | Category | n | % |
| [Continuous in years, as reported] | 39.0 | 18 | 90 | 18-25 (3) | 366 | 22.9 |
|  |  |  |  | 26-35 (2) | 445 | 27.8 |
|  |  |  |  | 36-50 (1) | 453 | 28.3 |
|  |  |  |  | 51-90 (0) | 335 | 20.9 |
|  |  |  |  | Don't know (.) | 1 | 00.1 |

*Respondent gender* [gender_dv]

| AB_R8-Q101. Respondent's gender | Original |  |  |  |  |
| --- | --- | --- | --- | --- | --- |
|  | n | % | Category | n | % |
| Male (1) | 799 | 49.9 | Male (1) | 799 | 49.9 |
| Female (2) | 801 | 50.1 | Female (0) | 801 | 50.1 |

*Respondent’s ‘race’* [race_dv]

*Respondent's 'race'* [race\_dv]
| AB_R8-Q102. Respondent’s race? | Original |  | Re-categorised categorical |  |  |
| --- | --- | --- | --- | --- | --- |
|  | n | % | Category | n | % |
| Black/African (1) | 1104 | 69.0 | Black/African (0) | 1104 | 69.0 |
| White/European (2) | 168 | 10.5 | White/European (1) | 168 | 10.5 |
| Coloured/Mixed race (3) | 244 | 15.3 | Coloured/Mixed race (2) | 244 | 15.3 |
| Arab/Lebanese/North Africa (4) | 0 | 0.0 |  |  |  |
| South Asian (Indian, Pakistani, etc.) (5) | 80 | 5.0 | South/East Asian (3) | 84 | 5.3 |
| East Asian (Chinese, Korean, Indonesian, etc.) (6) | 4 | 0.3 |  |  |  |
| Other [*Specify*] (9995) | 0 | 0.0 |  |  |  |
| Don’t know (9999) | 0 | 0.0 |  |  |  |

*Primary home language* [language_dv]

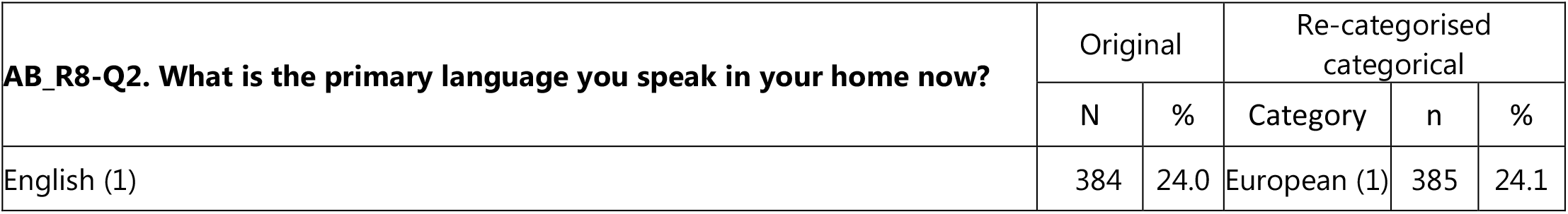
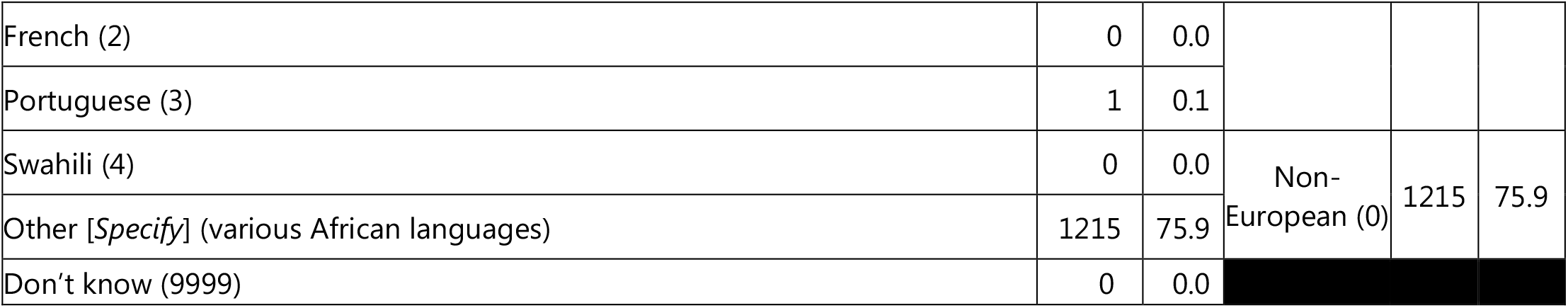

| AB_R8-Q2. What is the primary language you speak in your home now? | Original |  | Re-categorised categorical |  |  |
| --- | --- | --- | --- | --- | --- |
|  | N | % | Category | n | % |
| English (1) | 384 | 24.0 | European (1) | 385 | 24.1 |
| French (2) | 0 | 0.0 |  |  |  |
| Portuguese (3) | 1 | 0.1 |  |  |  |
| Swahili (4) | 0 | 0.0 | Non-European (0) | 1215 | 75.9 |
| Other [*Specify*] (various African languages) | 1215 | 75.9 |  |  |  |
| Don't know (9999) | 0 | 0.0 |  |  |  |

#### 2.2 Individual-level economic characteristics

*Educational attainment* [education_dv]

2.2 Individual-level economic characteristics *Educational attainment* [education\_dv]
| AB_R8-Q97. What is your highest level of education? | Original |  | Re-categorised ordinal |  |  |
| --- | --- | --- | --- | --- | --- |
|  | n | % | Category | n | % |
| No formal schooling (0) | 37 | 2.3 | Less than complete primary (4) | 164 | 10.2 |
| Informal schooling only (including Koranic schooling) (1) | 7 | 0.4 |  |  |  |
| Some primary schooling (2) | 120 | 7.5 |  |  |  |
| Primary school completed (3) | 79 | 4.9 | Complete primary but less than complete secondary (1) | 452 | 28.2 |
| Intermediate school or some secondary school / high school (4) | 373 | 23.3 |  |  |  |
| Secondary school / high school completed (5) | 545 | 34.1 | Secondary complete (0) | 545 | 34.1 |
| Post-secondary qualifications other than university, e.g. a diploma or degree from a polytechnic or college (6) | 175 | 10.9 | Non-university post-secondary (3) | 175 | 10.9 |
| Some university (7) | 90 | 5.6 | Some university or more (2) | 252 | 15.7 |
| University completed (8) | 127 | 7.9 |  |  |  |
| Post-graduate (9) | 35 | 2.2 |  |  |  |
| Refused or Don't know (98 or 99) | 12 | 0.8 | Refused/don't know (.) | 12 | 0.8 |

*Employment* [employ_dv]

| **AB_R8-Q95A. Do you have a job that pays a cash income? [If yes, ask:] Is it full time or part time? [If no, ask:] Are you currently looking for a job?** | Original |  | Re-categorised categorical |  |  |
| --- | --- | --- | --- | --- | --- |
|  | n | % | Category | n | % |
| No (not looking) (0) | 381 | 23.8 | No (not looking) (2) | 381 | 23.8 |
| No (looking) (1) | 608 | 38.0 | No (looking) (0) | 608 | 38.0 |
| Yes, part time (2) | 197 | 12.3 | Yes, part time (3) | 197 | 12.3 |
| Yes, full time (3) | 396 | 24.8 | Yes, full time (1) | 396 | 24.8 |
| Refused or Don't know (8 or 9) | 18 | 1.1 | Refused or Don't know (.) | 18 | 1.1 |

*Individual assets* [persradio_dv; perstv_dv; persvehicle_dv; perscomputer_dv; persbank_dv; persmobile_dv]

| **AB_R8-Q92. Which of these things do you personally own...?** | Original - Yes |  | Re-categorised categorical - Yes |  |  |
| --- | --- | --- | --- | --- | --- |
|  | n | % | Category | n | % |
| **92A.** Radio (Yes: 2; No: 1 or 0; Don't know: 9) | 1102 | 68.9 | Radio – Yes (1) | 1102 | 68.9 |
| **92B.** Television (Yes: 2; No: 1 or 0; Don't know: 9) | 1120 | 70.0 | TV – Yes (1) | 1120 | 70.0 |
| **92C.** Motor vehicle or motorcycle (Yes: 2; No: 1 or 0; Don't know: 9) | 453 | 28.3 | Vehicle – Yes (1) | 453 | 28.3 |
| **92D.** Computer (Yes: 2; No: 1 or 0; Don't know: 9) | 582 | 36.4 | Computer – Yes (1) | 582 | 36.4 |
| **92E.** Bank account (Yes: 2; No: 1 or 0; Don't know: 9) | 1305 | 81.6 | Account – Yes (1) | 1305 | 81.6 |
| **92F.** Mobile phone (Yes: 2; No: 1 or 0; Don't know: 9) | 1452 | 90.8 | Mobile – Yes (1) | 1452 | 90.8 |

#### 2.3 Household-level economic characteristics

*Dwelling structure* [dwelling_dv]

2.3 Household-level economic characteristics *Dwelling structure* [dwelling\_dv]
| **AB_R8-Q104. In what type of shelter does the respondent live?** | Original |  | Re-categorised |  |  |
| --- | --- | --- | --- | --- | --- |
|  | n | % | Category | n | % |
| Non-traditional/Formal house (1) | 1176 | 73.5 | Formal house/flat (0) | 1264 | 79.7 |
| Flat in block of flats (4) | 88 | 5.5 |  |  |  |
| Traditional house/Hut (2) | 119 | 7.4 | Single room or informal dwelling (1) | 322 | 20.3 |
| Temporary structure/Shack (3) | 150 | 9.4 |  |  |  |
| Single room in a larger dwelling structure or backyard (5) | 48 | 3.0 |  |  |  |
| Hostel in an industrial compound or farming compound (7) | 5 | 0.3 |  |  |  |
| Other (8) | 14 | 0.9 | Other (.) | 13 | 0.9 |

*Dwelling services, utilities and amenities – mains electricity* [hhldelectric_dv]

| **AB_R8-Q94A. Do you have an electric connection to your home from the [national power grid]?** | Original |  | Re-categorised categorical |  |  |
| --- | --- | --- | --- | --- | --- |
|  | n | % | Category | n | % |
| No (0) | 146 | 9.2 | No (1) | 146 | 9.2 |
| Yes (1) | 1446 | 90.4 | Yes (0) | 1446 | 90.4 |
| Refused to answer or Don't know (8; 9) | 8 | 0.5 | Refused or Don't know (.) | 8 | 0.5 |

*Dwelling services, utilities and amenties – water supply* [hhldwater_dv]

*Dwelling services, utilities and amenities – water supply [hhldwater\_dv]*
| **AB_R8-Q93A. What is your main source of water for household use?** | Original |  | Re-categorised categorical |  |  |
| --- | --- | --- | --- | --- | --- |
|  | n | % | Category | n | % |
| Piped water into dwelling (1) | 856 | 53.5 | Piped water into dwelling (0) | 856 | 53.5 |
| Piped water into yard, plot or compound (2) | 378 | 23.6 | Piped water into yard, plot or compound (1) | 378 | 23.6 |
| Public tap or standpipe (3) | 196 | 12.3 | Public tap or standpipe (2) | 196 | 12.3 |
| Tubewell or borehole (4) | 42 | 2.6 | Protected borehole, well or spring (3) | 55 | 3.4 |
| Protected dug well (5) | 6 | 0.4 |  |  |  |
| Protected spring (7) | 7 | 0.4 |  |  |  |
| Unprotected dug well (6) | 6 | 0.4 | Unprotected water source (4) | 40 | 2.6 |
| Unprotected spring (8) | 4 | 0.3 |  |  |  |
| Rainwater collection (9) | 10 | 0.6 |  |  |  |
| Surface water, like a river, dam, lake, pond, stream, canal or irrigation channel (13) | 20 | 1.3 |  |  |  |
| Bottled water (10) | 2 | 0.1 | Purchased water (5) | 49 | 3.0 |
| Purchased from a cart with a small tank or drum (11) | 15 | 0.9 |  |  |  |
| Purchased from a tanker truck (12) | 32 | 2.0 |  |  |  |
| Other (95) | 20 | 1.3 | Other, Refused or Don't know (.) | 26 | 1.6 |
| Refused (98) | 3 | 0.2 |  |  |  |
| Don't know (99) | 3 | 0.2 |  |  |  |

*Dwelling utilities, services and amenities – toilet facilities* [hhldtoilet_dv]

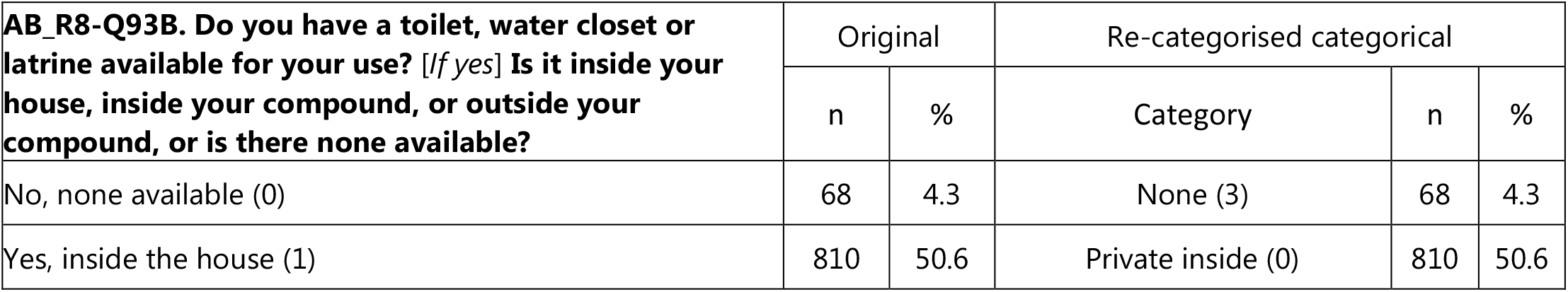
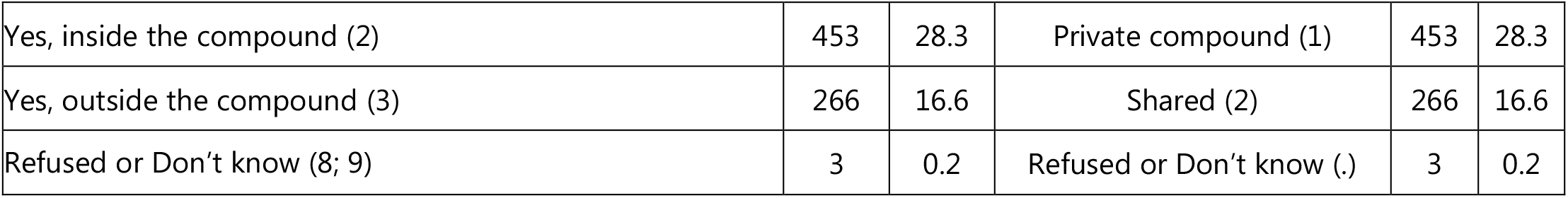

| **AB_R8-Q93B. Do you have a toilet, water closet or latrine available for your use? [If yes] Is it inside your house, inside your compound, or outside your compound, or is there none available?** | Original |  | Re-categorised categorical |  |  |
| --- | --- | --- | --- | --- | --- |
|  | n | % | Category | n | % |
| No, none available (0) | 68 | 4.3 | None (3) | 68 | 4.3 |
| Yes, inside the house (1) | 810 | 50.6 | Private inside (0) | 810 | 50.6 |
| Yes, inside the compound (2) | 453 | 28.3 | Private compound (1) | 453 | 28.3 |
| Yes, outside the compound (3) | 266 | 16.6 | Shared (2) | 266 | 16.6 |
| Refused or Don't know (8; 9) | 3 | 0.2 | Refused or Don't know (.) | 3 | 0.2 |

*Household assets* [hhldradio_dv; hhldtv_dv; hhldvehicle_dv; hhldcomputer_dv; hhldbank_dv; hhldmobile_dv]

*Household assets* [hhldradio\_dv; hhldtv\_dv; hhldvehicle\_dv; hhldcomputer\_dv; hhldbanks\_dv; hhldmobile\_dv]
| **AB_R8-Q92. Which of these things... does anyone [including respondent]... in the household own?** | Original - Yes |  | Re-categorised categorical - Yes |  |  |
| --- | --- | --- | --- | --- | --- |
|  | n | % | Category | n | % |
| **92A.** Radio (Yes: 1 or 2; No: 0; Don't know: 9) | 1360 | 85.0 | Radio – Yes (1) | 1360 | 85.0 |
| **92B.** Television (Yes: 1 or 2; No: 0; Don't know: 9) | 1464 | 91.5 | TV – Yes (1) | 1464 | 91.5 |
| **92C.** Motor vehicle or motorcycle (Yes: 1 or 2; No: 0; Don't know: 9) | 763 | 47.7 | Vehicle – Yes (1) | 763 | 47.7 |
| **92D.** Computer (Yes: 1 or 2; No: 0; Don't know: 9) | 774 | 48.4 | Computer – Yes (1) | 774 | 48.4 |
| **92E.** Bank account (Yes: 1 or 2; No: 0; Don't know: 9) | 1442 | 90.1 | Account – Yes (1) | 1442 | 90.1 |
| **92F.** Mobile phone (Yes: 1 or 2; No: 0; Don't know: 9) | 1529 | 95.6 | Mobile – Yes (1) | 1529 | 95.6 |

### Part 3: The distribution of specified exposures and outcomes, and selected covariates amongst South African respondents to the postponed Round 8 of the Afrobarometer survey *with* and *without* complete data on all variables

*Included vs. excluded respondents in the study’s analyses* [include_dv]

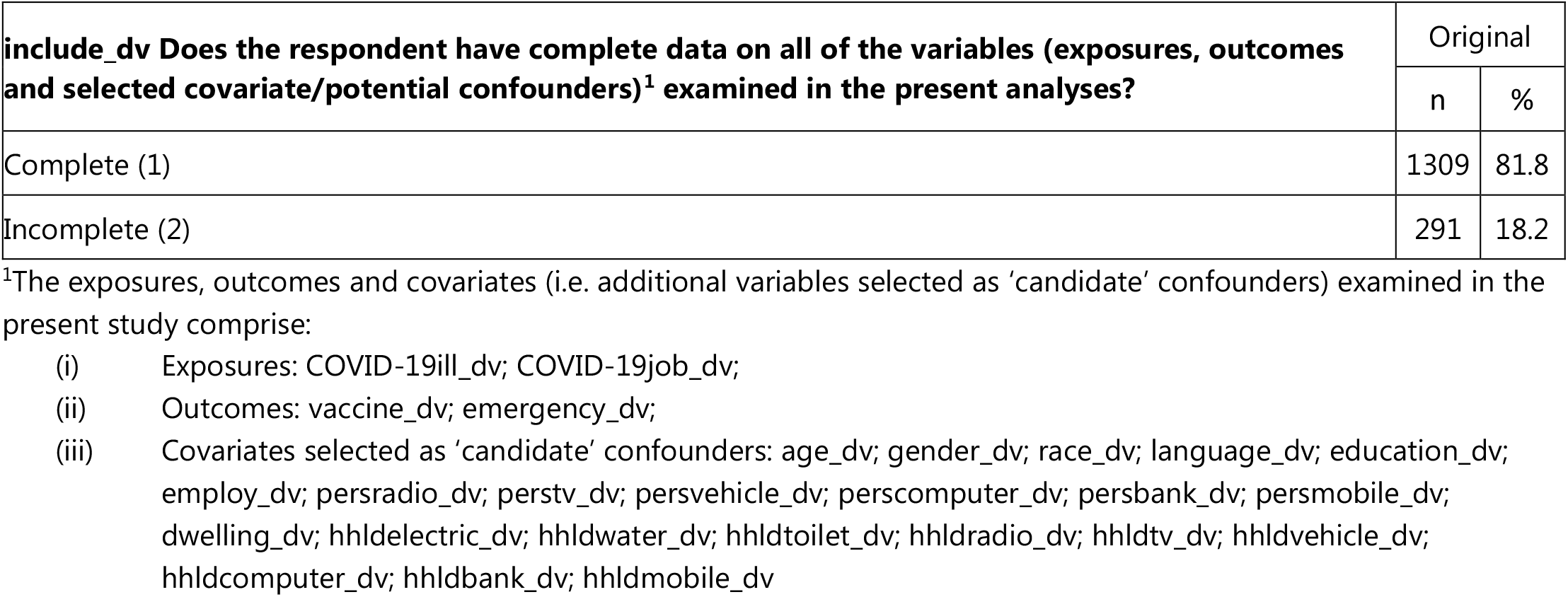

| **include_dv Does the respondent have complete data on all of the variables (exposures, outcomes and selected covariate/potential confounders)<sup>1</sup> examined in the present analyses?** | Original |  |
| --- | --- | --- |
|  | n | % |
| Complete (1) | 1309 | 81.8 |
| Incomplete (2) | 291 | 18.2 |

<sup>1</sup>The exposures, outcomes and covariates (i.e. additional variables selected as 'candidate' confounders) examined in the present study comprise:
- (i) Exposures: COVID-19ill\_dv; COVID-19job\_dv; - (ii) Outcomes: vaccine\_dv; emergency\_dv; - (iii) Covariates selected as 'candidate' confounders: age\_dv; gender\_dv; race\_dv; language\_dv; education\_dv; employ\_dv; persradio\_dv; perstv\_dv; persvehicle\_dv; perscomputer\_dv; persbank\_dv; persmobile\_dv; dwelling\_dv; hhldelectric\_dv; hhldwater\_dv; hhldtoilet\_dv; hhldradio\_dv; hhldtv\_dv; hhldvehicle\_dv; hhldcomputer\_dv; hhldbanks\_dv; hhldmobile\_dv

### Part 4: Multivariable statistical analyses

**Table S1.**
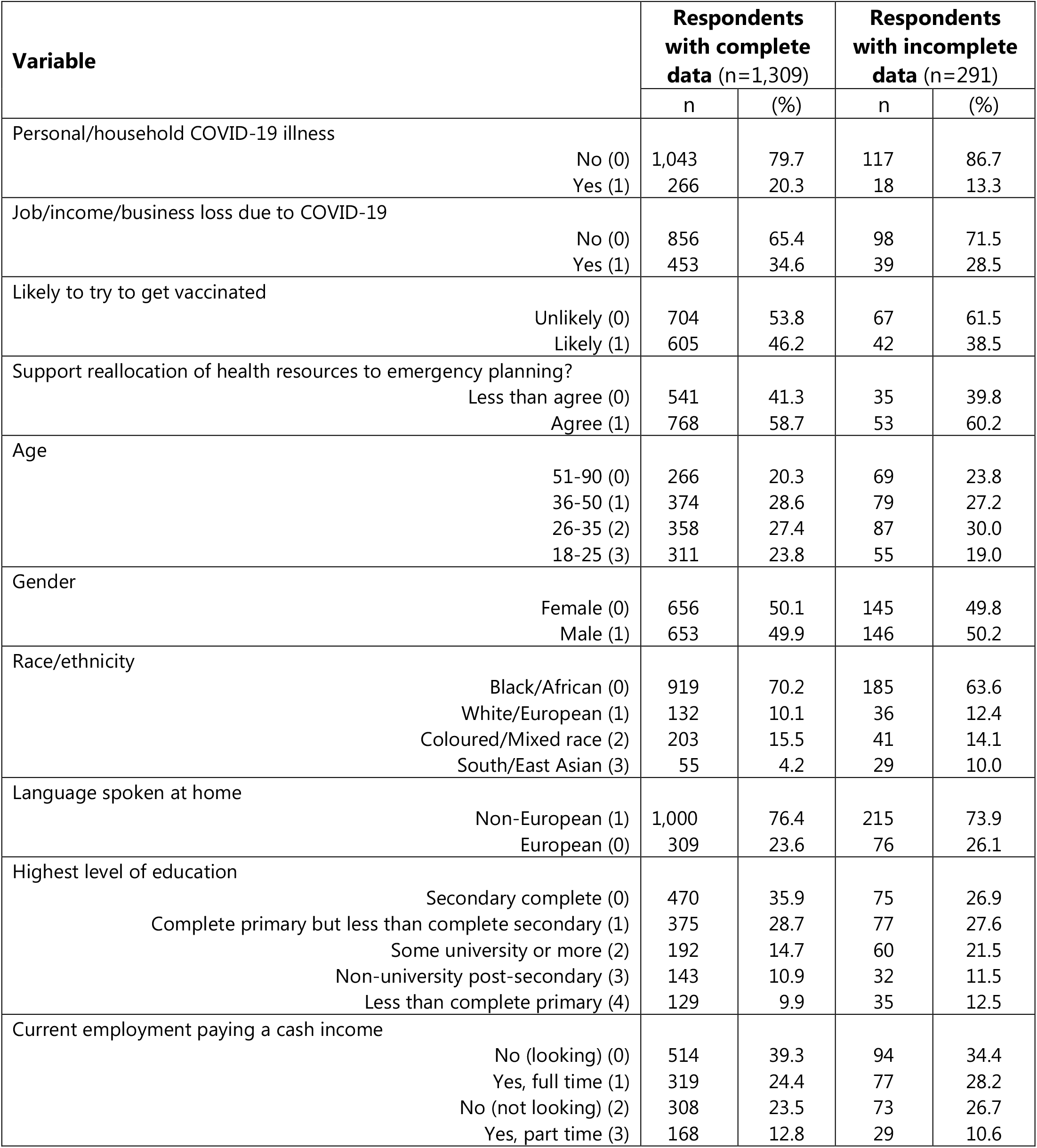
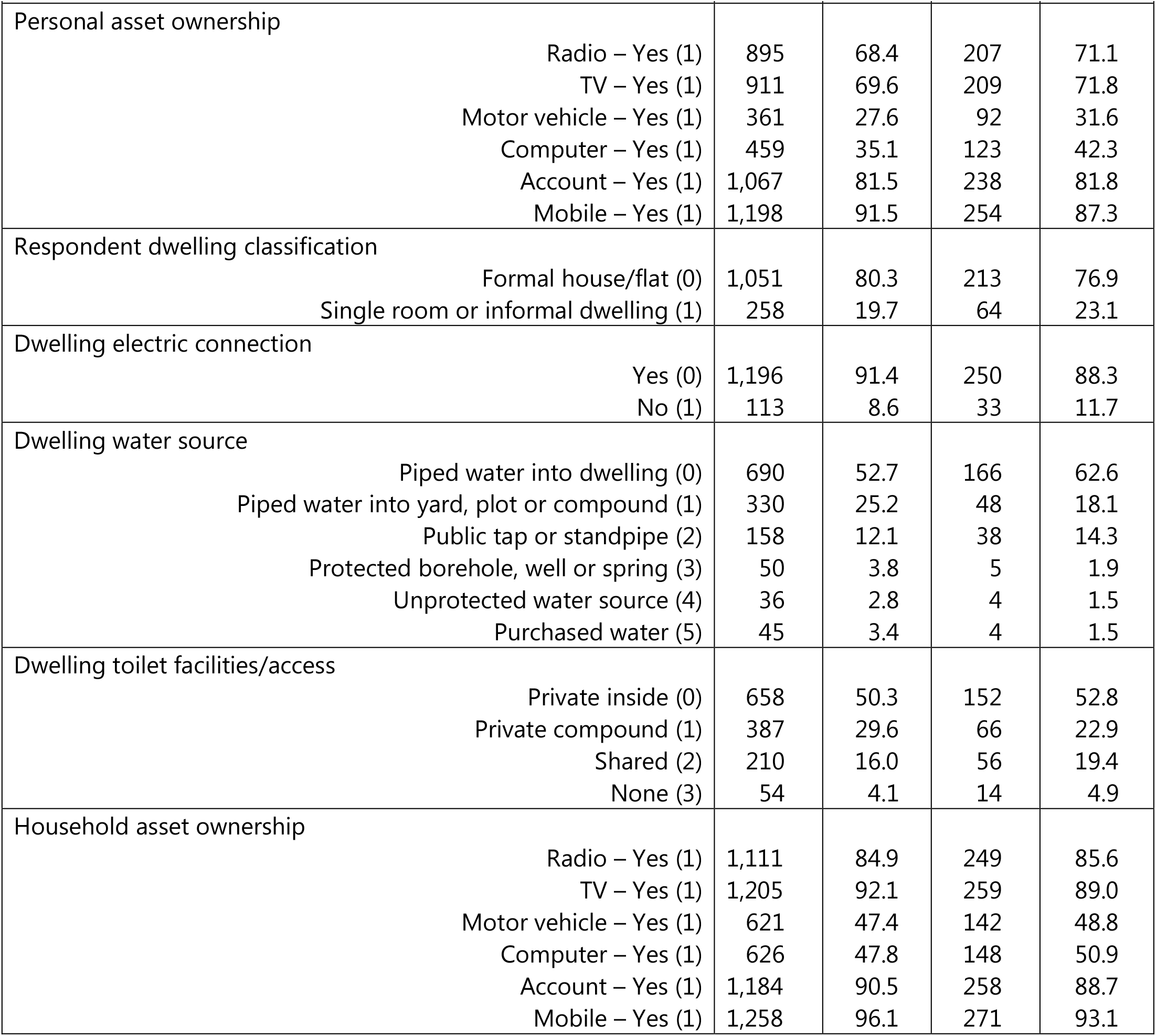
The distribution of exposure, outcome and ‘candidate’ confounder variables amongst: (i) the 1,309 South African respondents to Round 8 of the Afrobarometer surveys who provided answers to all 30 survey items (including the 2 specified exposures, 2 specified outcome and 26 ‘candidate’ confounder covariates), and (ii) the 291 respondents (18.2%) with missing data on one or more of these variables.

| Variable | Respondents with complete data (n=1,309) |  | Respondents with incomplete data (n=291) |  |
| --- | --- | --- | --- | --- |
|  | n | (%) | n | (%) |
| Personal/household COVID-19 illness |  |  |  |  |
| No (0) | 1,043 | 79.7 | 117 | 86.7 |
| Yes (1) | 266 | 20.3 | 18 | 13.3 |
| Job/income/business loss due to COVID-19 |  |  |  |  |
| No (0) | 856 | 65.4 | 98 | 71.5 |
| Yes (1) | 453 | 34.6 | 39 | 28.5 |
| Likely to try to get vaccinated |  |  |  |  |
| Unlikely (0) | 704 | 53.8 | 67 | 61.5 |
| Likely (1) | 605 | 46.2 | 42 | 38.5 |
| Support reallocation of health resources to emergency planning? |  |  |  |  |
| Less than agree (0) | 541 | 41.3 | 35 | 39.8 |
| Agree (1) | 768 | 58.7 | 53 | 60.2 |
| Age |  |  |  |  |
| 51-90 (0) | 266 | 20.3 | 69 | 23.8 |
| 36-50 (1) | 374 | 28.6 | 79 | 27.2 |
| 26-35 (2) | 358 | 27.4 | 87 | 30.0 |
| 18-25 (3) | 311 | 23.8 | 55 | 19.0 |
| Gender |  |  |  |  |
| Female (0) | 656 | 50.1 | 145 | 49.8 |
| Male (1) | 653 | 49.9 | 146 | 50.2 |
| Race/ethnicity |  |  |  |  |
| Black/African (0) | 919 | 70.2 | 185 | 63.6 |
| White/European (1) | 132 | 10.1 | 36 | 12.4 |
| Coloured/Mixed race (2) | 203 | 15.5 | 41 | 14.1 |
| South/East Asian (3) | 55 | 4.2 | 29 | 10.0 |
| Language spoken at home |  |  |  |  |
| Non-European (1) | 1,000 | 76.4 | 215 | 73.9 |
| European (0) | 309 | 23.6 | 76 | 26.1 |
| Highest level of education |  |  |  |  |
| Secondary complete (0) | 470 | 35.9 | 75 | 26.9 |
| Complete primary but less than complete secondary (1) | 375 | 28.7 | 77 | 27.6 |
| Some university or more (2) | 192 | 14.7 | 60 | 21.5 |
| Non-university post-secondary (3) | 143 | 10.9 | 32 | 11.5 |
| Less than complete primary (4) | 129 | 9.9 | 35 | 12.5 |
| Current employment paying a cash income |  |  |  |  |
| No (looking) (0) | 514 | 39.3 | 94 | 34.4 |
| Yes, full time (1) | 319 | 24.4 | 77 | 28.2 |
| No (not looking) (2) | 308 | 23.5 | 73 | 26.7 |
| Yes, part time (3) | 168 | 12.8 | 29 | 10.6 |
|  | n | (%) | n | (%) |
| Personal asset ownership |  |  |  |  |
| Radio – Yes (1) | 895 | 68.4 | 207 | 71.1 |
| TV – Yes (1) | 911 | 69.6 | 209 | 71.8 |
| Motor vehicle – Yes (1) | 361 | 27.6 | 92 | 31.6 |
| Computer – Yes (1) | 459 | 35.1 | 123 | 42.3 |
| Account – Yes (1) | 1,067 | 81.5 | 238 | 81.8 |
| Mobile – Yes (1) | 1,198 | 91.5 | 254 | 87.3 |
| Respondent dwelling classification |  |  |  |  |
| Formal house/flat (0) | 1,051 | 80.3 | 213 | 76.9 |
| Single room or informal dwelling (1) | 258 | 19.7 | 64 | 23.1 |
| Dwelling electric connection |  |  |  |  |
| Yes (0) | 1,196 | 91.4 | 250 | 88.3 |
| No (1) | 113 | 8.6 | 33 | 11.7 |
| Dwelling water source |  |  |  |  |
| Piped water into dwelling (0) | 690 | 52.7 | 166 | 62.6 |
| Piped water into yard, plot or compound (1) | 330 | 25.2 | 48 | 18.1 |
| Public tap or standpipe (2) | 158 | 12.1 | 38 | 14.3 |
| Protected borehole, well or spring (3) | 50 | 3.8 | 5 | 1.9 |
| Unprotected water source (4) | 36 | 2.8 | 4 | 1.5 |
| Purchased water (5) | 45 | 3.4 | 4 | 1.5 |
| Dwelling toilet facilities/access |  |  |  |  |
| Private inside (0) | 658 | 50.3 | 152 | 52.8 |
| Private compound (1) | 387 | 29.6 | 66 | 22.9 |
| Shared (2) | 210 | 16.0 | 56 | 19.4 |
| None (3) | 54 | 4.1 | 14 | 4.9 |
| Household asset ownership |  |  |  |  |
| Radio – Yes (1) | 1,111 | 84.9 | 249 | 85.6 |
| TV – Yes (1) | 1,205 | 92.1 | 259 | 89.0 |
| Motor vehicle – Yes (1) | 621 | 47.4 | 142 | 48.8 |
| Computer – Yes (1) | 626 | 47.8 | 148 | 50.9 |
| Account – Yes (1) | 1,184 | 90.5 | 258 | 88.7 |
| Mobile – Yes (1) | 1,258 | 96.1 | 271 | 93.1 |

**Table S2.** Logistic regression analyses exploring the relationship between each of the two specified exposures (E1: personal/household COVID-19 illness; and E2: job/income/business loss due to COVID-19) and each of the two specified outcomes (O1: likelihood of getting vaccinated; and O2: support for reallocation of health resources to emergency health planning). Separate models compared these relationships (E1→O1, E1→O2, E2→O1 and E2→O2) both before (Model 1) and after adjustment for n=9 (Model 2) and n=22 (Model 3) selected sociodemographic and economic covariates. The n=9 covariates included in Model 2 were all considered likely to have preceded both exposures; while the n=13 additional covariates included in Model 3 were considered susceptible to change following the onset of the COVID-19 pandemic (and hence mis-specification as *preceding* confounders rather than *subsequent* mediators or consequences of the outcome). All results are presented as odds ratios (OR) with 95% confidence intervals (95%CI) in parentheses.

| Specified outcome O1: Likely to get vaccinated – Likely (1) vs. Unlikely (0) |  |  |  |
| --- | --- | --- | --- |
| Specified exposures: | Size of covariate adjustment set<sup>1</sup> |  |  |
|  | Model 1: n=0 | Model 2: n=9 | Model 3: n=22 |
|  | OR (95%CI) | OR (95%CI) | OR (95%CI) |
| **E1: Personal/household COVID-19 illness?** |  |  |  |
| No (0) | Referent | Referent | Referent |
| Yes (1) | 1.04 (0.79,1.36) | 0.96 (0.72,1.28) | 0.98 (0.73,1.31) |
| **E2: Job/income/business loss due to COVID-19?** |  |  |  |
| No (0) | Referent | Referent | Referent |
| Yes (1) | 0.58 (0.47,0.75) | 0.60 (0.47,0.77) | 0.62 (0.48,0.79) |

| Specified outcome O2: Support health resource reallocation – Agree (1) vs. Less than agree (0) |  |  |  |
| --- | --- | --- | --- |
| Specified exposures: | Size of covariate adjustment set<sup>1</sup> |  |  |
|  | Model 4: n=0 | Model 5: n=9 | Model 6: n=22 |
|  | OR (95%CI) | OR (95%CI) | OR (95%CI) |
| **E1: Personal/household COVID-19 illness?** |  |  |  |
| No (0) | Referent | Referent | Referent |
| Yes (1) | 0.92 (0.70,1.21) | 0.95 (0.71,1.26) | 0.97 (0.73,1.30) |
| **E2: Job/income/business loss due to COVID-19?** |  |  |  |
| No (0) | Referent | Referent | Referent |
| Yes (1) | 0.99 (0.79,1.25) | 1.02 (0.80,1.30) | 1.05 (0.82,1.34) |

<sup>1</sup>Covariate adjustment sets for:
**Model 1 & 4** included n=0 covariates (i.e. no adjustment);
**Model 2 & 5** included n=9 covariates (comprising: age; gender; race/ethnicity; language; education; type of dwelling; and household services [electricity; water; and toilet facilities]); and
**Model 3 & 6** included n=22 covariates (comprising all n=9 of the covariates included in **Model 2 & 5**, in addition to: employment; and both personal and household ownership of n=6 assets [radio; tv; motor vehicle; computer; bank account; mobile phone]).

**Part 4: Figure S1.**
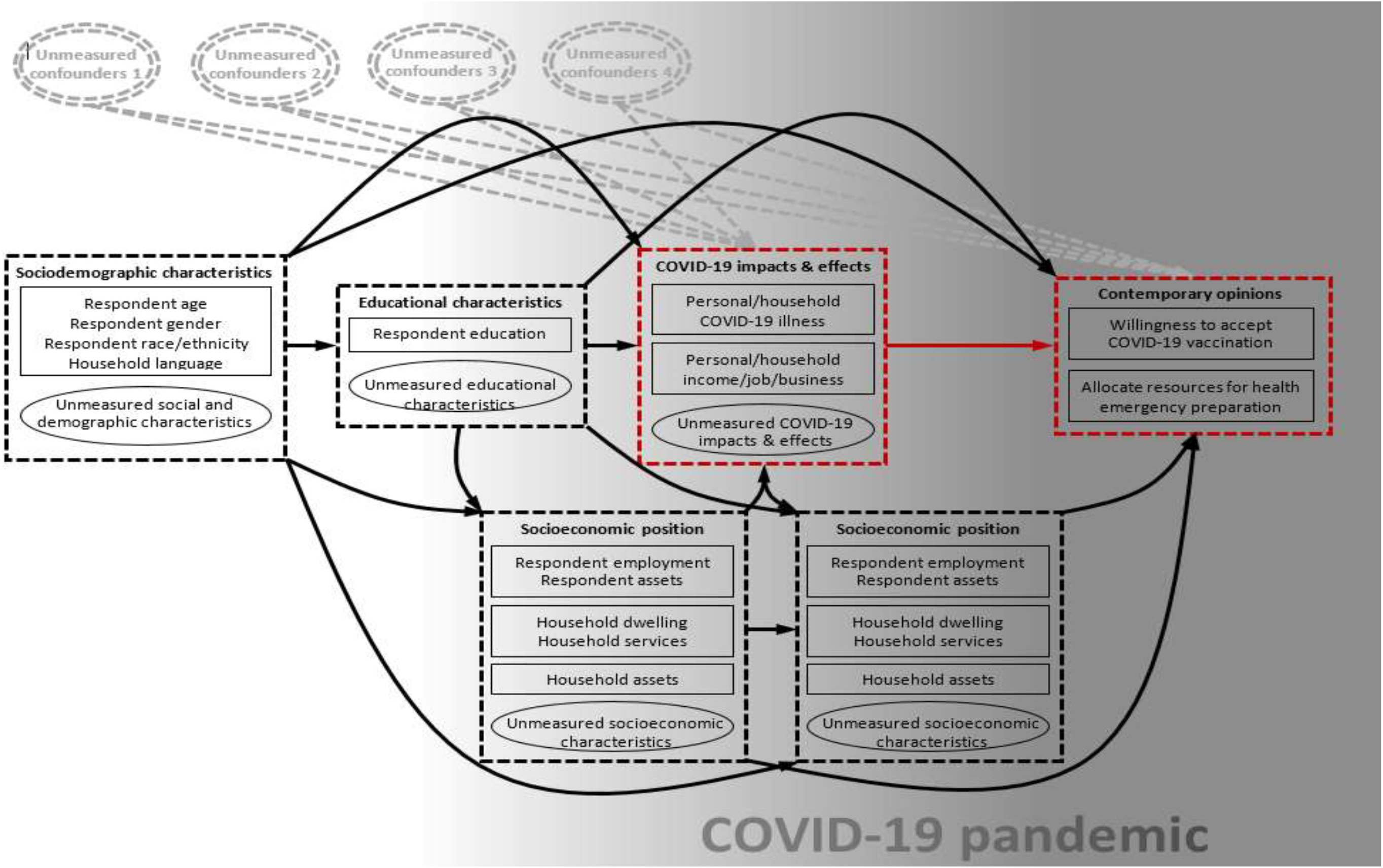
A theoretical causal path diagram, drawn in the form of a directed acyclic graph (DAG) in which each of the measured (rectangles) and unmeasured (ellipses) variables of relevance to the present study have been arranged in their hypothesised temporal sequence (from left to right), with each preceding variable assumed to act as a *probabilisitic* cause of all subsequent variables. The specified exposures and outcomes, and the focal relationships between these, have been indicated in red. Variables indicative of socioeconomic position are assumed to operate as time-variant covariates that may have crystallised (as/when surveyed) either before or after the onset of the COVID-19 pandemic. Examples of the many different unmeasured sets of covariates (indicated by ellipses with double outlines) likely to contribute confounder bias to estimates of the focal relationship examined in the present study, have been included to emphasise their potential impact.

